# A Novel Multiplex PCR Based Detection Assay Using Saliva or Nasopharyngeal Samples for SARS-Cov-2, Influenza A and B – Clinical Validation and Utility for Mass Surveillance

**DOI:** 10.1101/2021.01.13.21249629

**Authors:** Nikhil S Sahajpal, Ashis K Mondal, Sudha Ananth, Allan Njau, Pankaj Ahluwalia, Eesha Oza, Ted M Ross, Vamsi Kota, Arvind Kothandaraman, Sadanand Fulzele, Madhuri Hegde, Alka Chaubey, Amyn M Rojiani, Ravindra Kolhe

**Affiliations:** Department of Pathology, Medical College of Georgia, Augusta University, GA, U.S.A.; Department of Pathology, Aga Khan University Hospital, Nairobi, Kenya; Center for Vaccines and Immunology, University of Georgia, GA, U.S.A.; Department of Medicine, Medical College of Georgia, Augusta University, GA, U.S.A.; Center for Healthy Aging, Medical College of Georgia, Augusta University, Augusta, GA; Global Laboratory Services, Perkin Elmer, Waltham, U.S.A.

**Author notes:** Corresponding author Ravindra Kolhe MD, PhD., Medical College of Georgia| Augusta University., BAE 2576, 1120 15th Street| Augusta, GA 30912.

## Abstract

**Background:** The COVID-19 pandemic has resulted in a significant diversion of human and material resources to COVID-19 diagnostics, to the extent that testing of viral pathogens normally contributing to seasonal respiratory tract infections have been markedly neglected. The global health burden due to influenza viruses and co-infection in COVID-19 patients remains undocumented but clearly pose serious public health consequences. To address these clinical and technical challenges, we have optimized and validated a highly sensitive RT-PCR based multiplex assay for the detection of SARS-CoV-2, Influenza A and B viruses in a single test.

**Methods:** This study evaluated clinical specimens (n=1411) that included 1019 saliva and 392 nasopharyngeal swab (NPS) samples collected in either healthcare or community setting. Samples were tested using two assays: FDA-EUA approved SARS-CoV-2 assay that targets *N* and *ORF1ab* gene, and the PKamp RT-PCR based assay that targets SARS-CoV-2, Influenza viruses A and B. The limit of detection (LoD) studies was conducted as per the FDA guidelines using SARS-CoV-2 and Influenza A and B reference control materials.

**Results:** Of the 1019 saliva samples, 17.0% (174/1019) tested positive for SARS-CoV-2 using either assay. The detection rate for SARS-CoV-2 was higher with our multiplex assay compared to SARS-specific assay [91.9% (160/174) vs. 87.9% (153/174)], respectively. Of the 392 NPS samples, 10.4% (41/392) tested positive for SARS-CoV-2 using either assay. The detection rate for SARS-CoV-2 was higher with our multiplex assay compared to SARS-specific assay [97.5% (40/41) vs. 92.1% (39/41)], respectively. The Ct values for SARS-CoV-2 were comparable between the two assays, whereas the Ct values of the housekeeping gene was significantly lower with multiplex assay compared to SARS-specific assay. The LoD was established as 60 copies/ml for SARS-CoV-2 and 180 copies/ml for Influenza A and B viruses for both saliva and NPS samples.

**Conclusion:** This study presents clinical validation of a multiplex PCR assay for testing SARS-CoV-2, Influenza A and B viruses, using NPS and saliva samples, and demonstrates the feasibility of implementing the assay without disrupting the existing laboratory workflow. This novel assay uses the same instruments, sample types, supplies, and laboratory personnel as needed for the testing of SARS-CoV-2 virus.

## Introduction

The outbreak of COVID-19 (caused by SARS-CoV-2) is currently a raging pandemic and has led to major socio-economic disruption worldwide. Since the identification of SARS-CoV-2 in the region of Wuhan, China, 77,343,652 confirmed cases with over 1,702,293 COVID-19 related deaths have been reported globally (https://coronavirus.jhu.edu/map.html, last accessed December 21, 2020). In an attempt to curtail the spread of the disease, testing for SARS-CoV-2 has been identified as the single most important measure. Implementation of testing for the virus in the USA and around the world has been facilitated by substantial emergency policy changes as well as state-sponsored funding [1, 2]. However, due to the diversion of personnel, resources, and supplies to SARS-CoV-2 testing, the testing of viral pathogens that normally cause seasonal respiratory tract infections has been marginalized. Acute respiratory tract infections (ARTIs) remain the leading cause of morbidity and mortality from infectious diseases. The Global Burden of Disease (2017) data has demonstrated that influenza contributed 11.5% of the total lower respiratory tract infections (LRTIs), leading to over 9 million hospitalizations and 145,000 deaths across all age groups in a single calendar year [3]. Further, co-infection with bacterial, fungal, or viral pathogens has been associated with disease severity and death in the current pandemic. A report originating from Wuhan, China, identified that 80% of COVID-19 patients had co-infection with at least one respiratory pathogen, with the most common being influenza viruses A and B (60 % and 53.30% respectively) [4]. The neglect of these co-circulating pathogens, especially influenza A and B viruses, has generated serious public health gaps both at a clinical and epidemiological level. The potential impact of co-circulating respiratory viral pathogens during the ongoing influenza season is of major concern at both local and international public health levels. Further, with the COVID-19 vaccination already in use, testing for pathogens, especially the influenza viruses in addition to SARS-CoV-2, is already becoming the next diagnostic testing emergency.

We must remain cognizant of the fact that the pandemic has caused a significant change in routine clinical diagnostic laboratories at the level of re-directing workflow, workforce, supplies, and validating new diagnostic tests [5]. It is only in the last few months that laboratories have streamlined testing for SARS-CoV-2, and therefore shifting to a new workflow and implementing new diagnostic tests, would only unsettle the already exhausted laboratory staff. To address these clinical and technical challenges, we have optimized and validated a highly sensitive RT-PCR based multiplex assay for the detection of SARS-CoV-2, Influenza A and B viruses in a single test, using the same workflow, instruments, sample types, supplies, and laboratory personnel currently utilized for SARS-CoV-2 testing. Additionally, the assay was validated for both nasopharyngeal swab (NPS) and saliva samples and demonstrated higher sensitivity in detecting SARS-CoV-2 virus compared to the FDA-EUA comparator assay (SARS-CoV-2 detection only). We contend that this novel multiplex assay can be used to screen for influenza viruses along with the detection of SARS-CoV-2 and can be readily implemented in laboratories across the globe, using essentially the same workflow and rapid turnaround time (TAT) (**Figure 1**).

**Figure 1.**
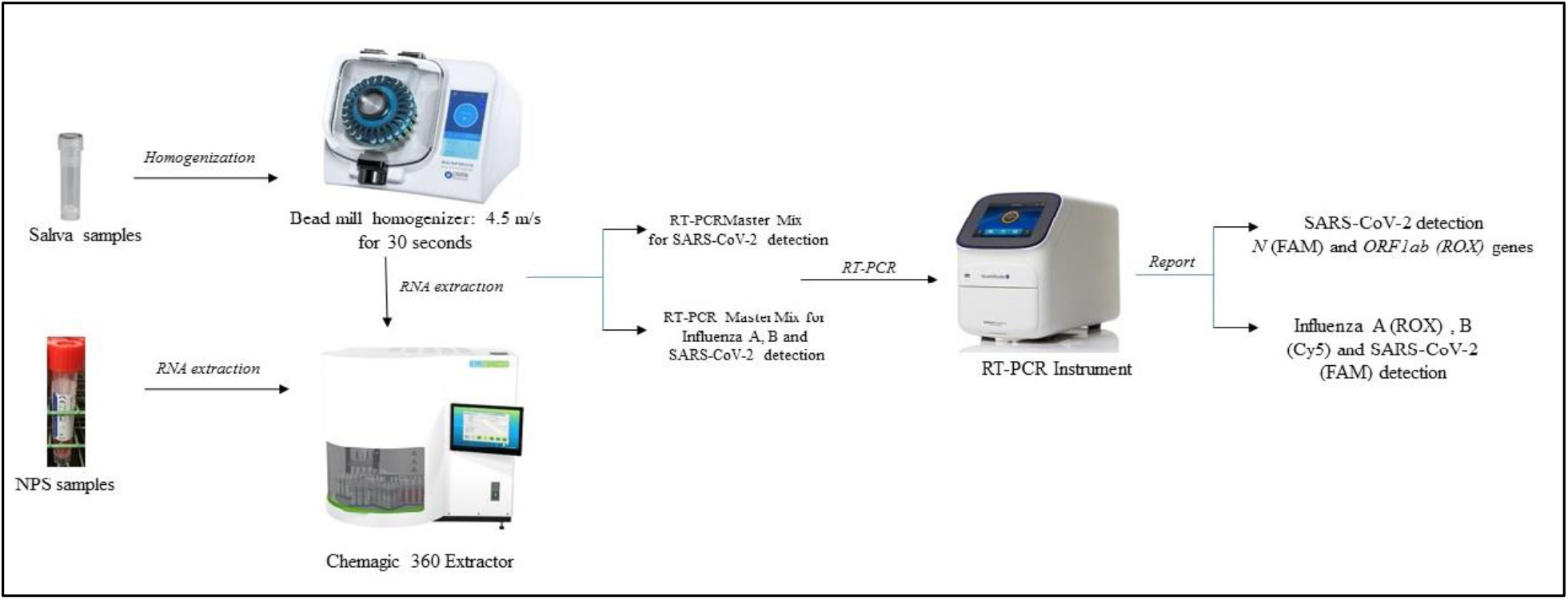
Schematic overview of sample processing steps for saliva and NPS samples for SARS-specific assay and the multiplex assay. The two assays differ in the preparation of RT-PCR master mix preparation, and the entire protocol remains the same.

## Material and Methods

### Study site and ethics

This single-center diagnostic study was conducted at Augusta University, GA, USA. This site is a Clinical Laboratory Improvement Amendments (CLIA) accredited laboratory for high complexity testing and is one of the main SARS-CoV-2 testing centers in the State of Georgia. The samples were processed under an approved HAC by the IRB Committee A (IRB REGISTRATION # 611298), Augusta University, GA. Based on the IRB approval, all PHI was removed and all data was anonymized before accessing for the study.

### Patient Specimens and setting

The study evaluated 1411 clinical specimens that included 1019 saliva and 392 NPS samples collected in either healthcare or community setting, tested using both the FDA-EUA approved SARS-CoV-2 and the PKamp RT-PCR based assays. As a standard protocol, NPS samples were collected by a healthcare worker using a sterile flocked swab placed in a sterile tube containing the viral transport medium (VTM) (Becton Dickinson, USA, cat no. 22053). The saliva samples were collected under the supervision of a healthcare worker in Omni tubes (Omni International, USA, SKU: 19-628D) without adding any media. All samples were stored at 4°C temperature and transported to the SARS-CoV-2 testing facility at Augusta University, GA, within 24 hours of sample collection, for further processing.

### FDA-EUA approved assay for the detection of SARS-CoV-2 (SARS-specific assay)

The assay is based on nucleic acid extraction followed by TaqMan-based RT-PCR assay to conduct *in vitro* transcription of SARS-CoV-2 RNA, DNA amplification, and fluorescence detection (FDA-EUA assay by PerkinElmer Inc. Waltham, USA). The assay targets specific genomic regions of the SARS-CoV-2: nucleocapsid (*N*) gene and *ORF1ab*. The TaqMan probes for the two amplicons are labeled with FAM and ROX fluorescent dyes, respectively, to generate target-specific signals. The assay includes an RNA internal control (IC, bacteriophage MS2) to monitor the processes from nucleic acid extraction to fluorescence detection. The IC probe is labeled with VIC fluorescent dye to differentiate its fluorescent signal from SARS-CoV-2 targets. The samples were resulted as positive or negative based on the Ct values specified by the manufacturer. For a detailed method, please refer to Sahajpal, NS, et al. [6].

### PKamp assay for the detection of SARS-CoV-2, Influenza A and B viruses (Multiplex assay)

The PKamp Respiratory SARS-CoV-2 RT-PCR Panel assay (PerkinElmer Inc. Waltham, USA) is a real-time reverse transcription-polymerase chain reaction (RT -PCR) multiplexed test intended for the simultaneous qualitative detection and differentiation of SARS-CoV-2, influenza A, influenza B, and respiratory syncytial virus (RSV). The oligonucleotide primers and probes for the detection of SARS-CoV-2 include the virus nucleocapsid (*N*) gene and *ORF1ab* gene. The primers and probes for the detection of influenza A and RSV target regions of matrix protein. The primers and probes for the detection of influenza B target the regions of the nuclear export protein (*NEP*) and nonstructural protein 1 (*NS1*) genes. An additional primer/probe set to detect the endogenous control targets the *RNase P* gene and is included in the test. Qualitative assessment is based on fluorescence detections with TaqMan probes labeled as SARS-CoV-2 (FAM), Influenza A (ROX), Influenza B (Cy5), Respiratory syncytial virus (Cy5.5), and RNase P (HEX/VIC).

### Protocol

In brief, all samples were vortexed, and an aliquot of 300µl from each sample (NPS or Saliva), as well as positive and negative controls, were added to respective wells in a 96 well plate. To each well, 4µl Poly(A) RNA, 10µl proteinase K, and 300µl lysis buffer 1 were added. The plate was placed on a semi-automated instrument (Chemagic 360 instrument, PerkinElmer Inc.) following the manufacturer’s protocol. Nucleic acid was extracted in a 96 well plate, with an elution volume of 60µl. From the extraction plate, 10µl of extracted nucleic acid and 5µl of PCR master mix was added to the respective wells in a 96 well PCR plate. The PCR method was set up as per the manufacturer’s protocol on Quantstudio 3 or 5 (Thermo Fisher Scientific, USA). The samples were resulted as positive or negative, based on the Ct values specified by the manufacturer (Supplementary file 1). Saliva samples were processed using the SalivaAll protocol [7].

### Limit of detection studies

The limit of detection (LoD) studies were conducted as per the FDA guidelines (https://www.fda.gov/medical-devices/coronavirus-disease-2019-covid-19-emergency-use-authorizations-medical-devices/vitro-diagnostics-euas). Briefly, SARS-CoV-2 reference control material (SARS-CoV-2: SeraCare (Mat. No. 0505-0159; Influenza A and B: SeraCare (Mat. No. 0515-0001) was spiked into the negative saliva and NPS samples to serve as positive samples at 540 copies/ml, 180 copies/ml, 60 copies/ml and 20 copies/ml concentrations. The lowest concentration detected in all three triplicates was determined as the preliminary LoD. To confirm the LoD, 20 replicates of preliminary LoD were analyzed and deemed as confirmed if at least 19/20 replicates were detected.

## Data Analysis

Data were analyzed for descriptive statistics and presented as a number (%) for categorical variables and mean ± standard deviation (SD) for continuous variables. Ct values were compared using Paired T-test.

## Results

### Clinical performance: Saliva samples

Of the 1019 saliva samples, 17.0% (174/1019) tested positive for SARS-CoV-2 using either assay. The detection rate for SARS-CoV-2 was higher with multiplex assay compared to SARS-specific assay [91.9% (160/174) vs. 87.9% (153/174)], respectively. The concordance for positive results between the two tests was 80.4% (140/174) (**Table 1**). The Ct values for SARS-CoV-2 were comparable between the two assays (SARS-CoV-2: 29.2 ± 7.2 vs. N: 29.8 ± 6.4, ORF1ab: 28.0 ± 7.1), whereas the Ct values of housekeeping gene was significantly lower with multiplex assay compared to SARS-specific assay [RNaseP: 20.6 ± 2.05 vs. IC: 32.5 ± 2.0 (p<0.001)], respectively (**Figure 2**). Further, the number of tests resulting as invalid was significantly less with the multiplex assay compared to the SARS-specific assay [1.7% (18/1019) vs. 5.6% (58/1019) (p< 0.001)].

**Table 1.** The overall and assay specific detection rate (%) and concordance data between two assays for saliva samples.

| Parameters | Total positive | Positive rate (%) |
| --- | --- | --- |
| Saliva sample (Both assays) | 174/1019 | 17 |
| Saliva (FDA-EUA assay) | 153/174 | 87.9 |
| Saliva (Multiplex assay) | 160/174 | 91.9 |
| Concordance | 140/174 | 80.4 |

**Figure 2.**
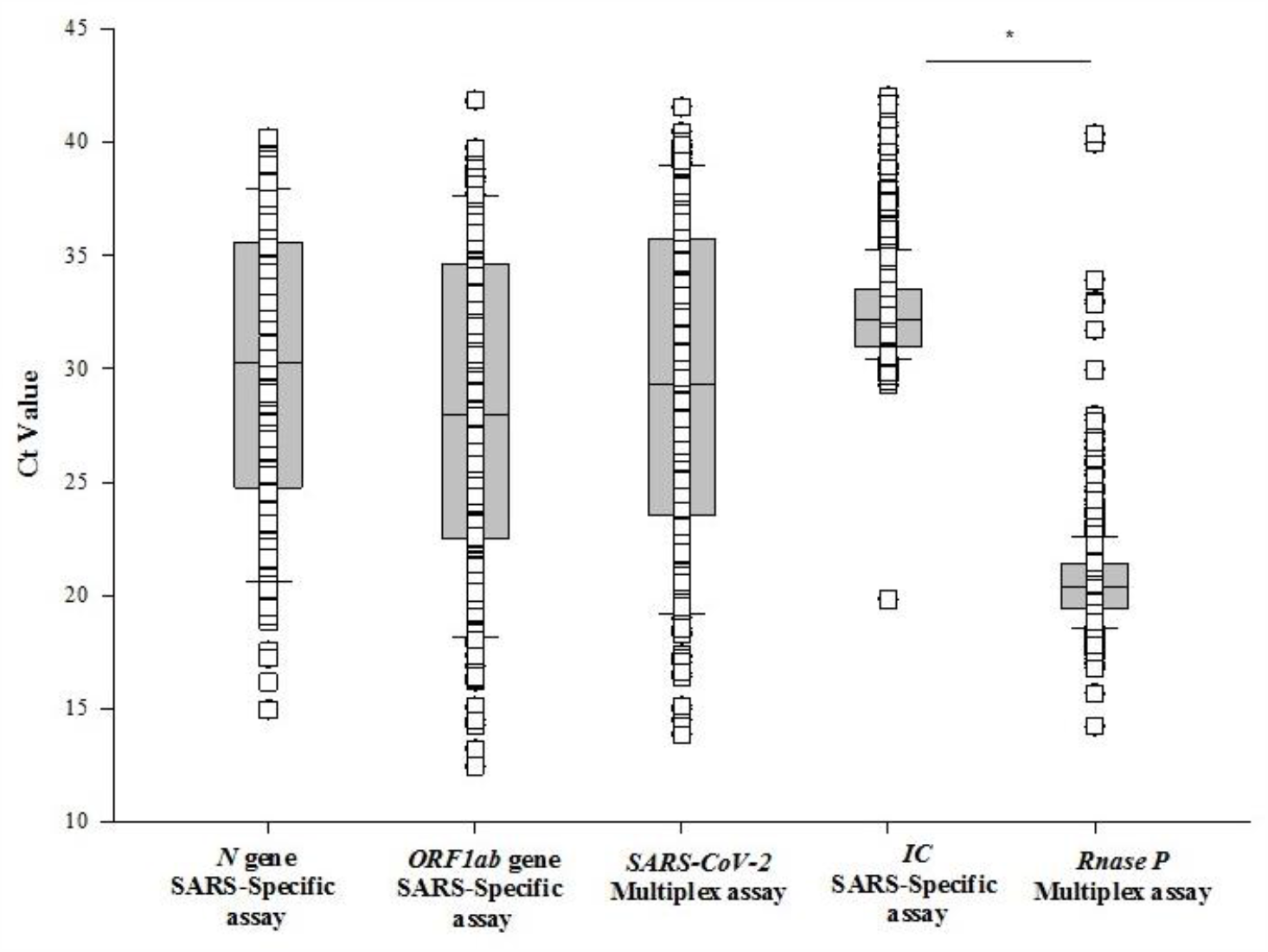
Plot demonstrating the Ct values of *N* and *ORF1ab* genes in SARS-specific assay compared to SARS-CoV-2 in the multiplex assay. Comparison of Ct values of housekeeping genes *IC* and *Rnase P* in SARS-specific and Multiplex assay, respectively, for saliva samples.

### Clinical performance: NPS samples

Of the 392 saliva samples, 10.4% (41/392) tested positive for SARS-CoV-2 using either assay. The detection rate for SARS-CoV-2 was higher with our multiplex assay compared to the SARS-specific assay [97.5% (40/41) vs. 95.1% (39/41)], respectively. The concordance for positive results between the two tests was 92.6% (38/41) (**Table 2**). The Ct values for SARS-CoV-2 were comparable between the two assays (SARS-CoV-2: 30.8 ± 8.8 vs. *N*: 31.3 ± 8.6, *ORF1ab*: 28.8 ± 8.6), whereas the Ct values of housekeeping gene was significantly lower with multiplex assay compared to SARS-specific assay [*RNaseP*: 23.1 ± 1.77 vs. *IC*: 31.3 ± 1.5 (p<0.001)], respectively (**Figure 3**).

**Table 2.** The overall and assay specific detection rate (%) and concordance data between two assays for NPS samples.

| Parameters | Total positive | Positive rate (%) |
| --- | --- | --- |
| NPS sample (Both assays) | 41/392 | 10.4 |
| NPS (SARS-specific assay) | 39/41 | 95.1 |
| NPS (Multiplex assay) | 40/41 | 97.5 |
| Concordance | 38/41 | 92.6 |

**Figure 3.**
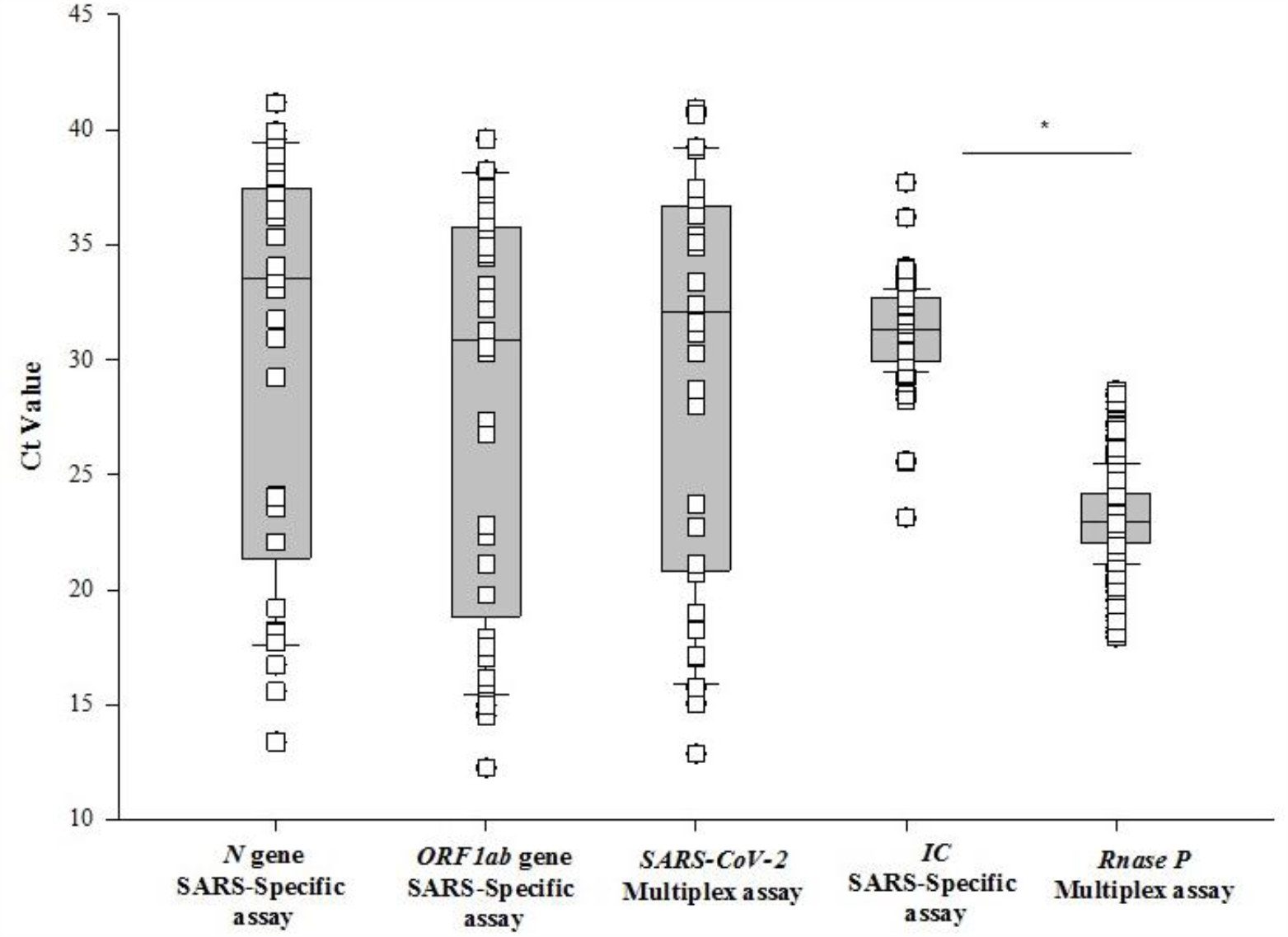
Plot demonstrating the Ct values of *N* and *ORF1ab* genes in SARS-specific assay compared to SARS-CoV-2 in the multiplex assay. Comparison of Ct values of housekeeping genes *IC* and *Rnase P* in SARS-specific and Multiplex assay, respectively, for NPS samples.

### Limit of detection studies: Saliva samples

In the preliminary LoD study using saliva samples, all replicates were detected at 60, 180, and 540 copies/ml for SARS-CoV-2 and the LoD was determined at 60 copies/ml, with 20/20 replicates being detected. The preliminary LoD evaluation for Influenza A and B identified all replicates at the four concentrations for influenza B, whereas all replicates at 180 and 540 copies/ml were positive for influenza B. The LoD was established at 180 copies/ml for both Influenza A, and B viruses, with 20/20 replicates being detected (**Table 3**).

**Table 3.** Limit of detection for SARS-CoV-2, Influenza A and B viruses for saliva samples.

| Preliminary LoD study |  | Saliva |  |
| --- | --- | --- | --- |
| Concentrations | SARS-CoV-2 (Ct/replicates) | Influenza A (Ct/replicates) | Influenza B (Ct/replicates) |
| 20 copies/ml | 37.0 ± 0.87 (2/3) | – (0/3) | 34.9 ± 1.29 (3/3) |
| 60 copies/ml | 34.7 ± 0.52 (3/3) | 37.1 (1/3) | 34.6 ± 0.61 (3/3) |
| 180 copies/ml | 33.2 ± 0.22 (3/3) | 35.7 ± 0.88 (3/3) | 32.3 ± 0.57 (3/3) |
| 540 copie/ml | 31.6 ± 0.21 (3/3) | 34.5 ± 1.24 (3/3) | 31.3 ± 0.55 (3/3) |
| LoD Confirmation | 60 copies/ml: 20/20 replicates detected | 180 copies/ml: 20/20 replicates detected | 180 copies/ml: 20/20 replicates detected |

### Limit of detection studies: NPS samples

In the preliminary LoD study using NPS samples, all replicates at the four tested concentrations (20, 60, 180, and 540 copies/ml) were detected for SARS-CoV-2, and the LoD was determined at 60 copies/ml, with 20/20 replicates being detected. The preliminary LoD evaluation for Influenza A and B identified all replicated at the four concentrations for influenza B, whereas all replicates at 180 and 540 copies/ml were detected for influenza B. The LoD was established at 180 copies/ml for both Influenza A, and B viruses, with 20/20 replicates being detected (**Table 4**).

**Table 4.**
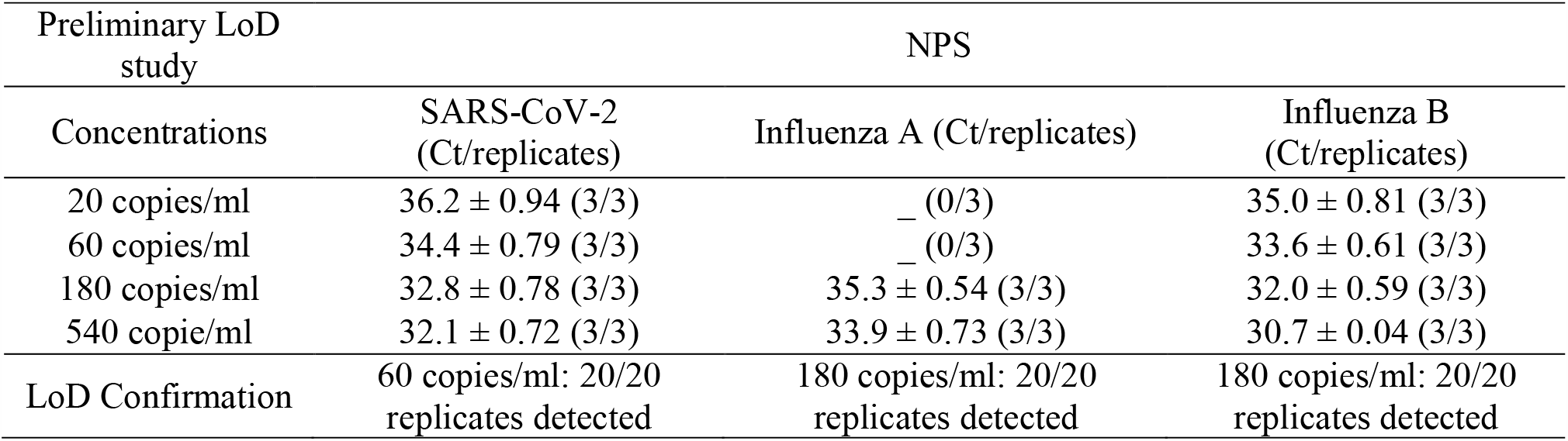
Limit of detection for SARS-CoV-2, Influenza A and B viruses for NPS samples.

| Preliminary LoD study |  | NPS |  |
| --- | --- | --- | --- |
| Concentrations | SARS-CoV-2 (Ct/replicates) | Influenza A (Ct/replicates) | Influenza B (Ct/replicates) |
| 20 copies/ml | 36.2 ± 0.94 (3/3) | – (0/3) | 35.0 ± 0.81 (3/3) |
| 60 copies/ml | 34.4 ± 0.79 (3/3) | – (0/3) | 33.6 ± 0.61 (3/3) |
| 180 copies/ml | 32.8 ± 0.78 (3/3) | 35.3 ± 0.54 (3/3) | 32.0 ± 0.59 (3/3) |
| 540 copie/ml | 32.1 ± 0.72 (3/3) | 33.9 ± 0.73 (3/3) | 30.7 ± 0.04 (3/3) |
| LoD Confirmation | 60 copies/ml: 20/20 replicates detected | 180 copies/ml: 20/20 replicates detected | 180 copies/ml: 20/20 replicates detected |

## Discussion

Diagnostic testing for COVID-19 has been executed in mammoth proportions worldwide, and this collective effort has helped frame regional policies for businesses, hospital preparedness, and defining public safety measures in a collective struggle to contain the spread of the disease [1, 2]. The emergent situation combined with limited knowledge in the early phase of the pandemic led to drastic changes in clinical diagnostic laboratories worldwide, requiring major workflow changes and diversion of supplies and workforce towards COVID-19 testing. The emergency continues to this day as laboratories work tirelessly to cope with the unprecedented sample volume and the rapidly evolving scenarios of the pandemic. Unfortunately, this has led to significant neglect of other circulating pathogens in the population, especially influenza viruses that typically have a substantial annual impact on global health [3, 8]. This relatively undocumented health loss due to seasonal influenza viruses has posed severe public health deficiencies, both at a clinical and epidemiological level [9, 10]. Further, poorly documented co-infections in patients with COVID-19 continue to amplify the morbidity and mortality associated with the pandemic [4, 11].

In an attempt to address these clinical and technical challenges, we propose a simplified protocol for the detection of SARS-CoV-2 along with common co-infections. We have optimized and validated a highly sensitive, RT-PCR based multiplex assay for detecting SARS-CoV-2, Influenza A, and B viruses in a single test, using the same workflow, instruments, sample types, supplies, and laboratory personnel needed for the testing of SARS-CoV-2 virus. The clinical evaluation of 1411 specimens that included 1019 saliva samples and 392 NPS samples demonstrated comparable performance of the multiplex assay to the SARS-specific FDA-EUA assay. Of the 1019 saliva samples evaluated with both assays, the detection rate was higher with the proposed multiplex assay compared to the SARS-specific assay (91.9% (160/174) vs. 87.9% (153/174). Ct values for SARS-CoV-2 with the multiplex assay were comparable to *N* and *ORF1ab* genes with the SARS-specific assay. The marginally higher performance of the multiplex assay compared to the SARS-specific assay might be attributed to the fact that in the multiplex assay, the amplification from two regions of the SARS-CoV-2 genome (*N* and *ORF1ab* genes) are detected with a single dye (FAM) for fluorescence detection. Thus, very weak positive samples that do not amplify sufficiently for the respective targets individually (*N*: FAM, *ORF1ab*: ROX) in the SARS-specific assay are being amplified to cross the threshold collectively in the multiplex assay [SARS-CoV-2 (*N, ORF1ab*): FAM). The concordance between the two assays was found to be 80.4% (140/174). Upon manual inspection of the Ct values, it was found that the discrepant results between the assays emerged for samples that show low viral loads with high Ct values, and only one gene detected (*N* or *ORF1ab*) with the SARS-specific assay. The findings were similar with NPS samples, with a slightly higher detection rate with multiplex assay compared to SARS-specific assay [97.5% (40/41) vs. 95.1% (39/41)], with a concordance of 92.6% (38/41) between the two assays. The Ct values for SARS-CoV-2 with the multiplex assay were comparable to *N* and *ORF1ab* gene with the SARS-specific assay. Further, the LoD studies performed with the SeraCare reference material demonstrated high sensitivity of the assay with LoD of 60 copies/ml for SARS-CoV-2 and 180 copies/ml for influenza A and B viruses, for both saliva and NPS samples. Notably, no samples resulted positive for Influenza viruses A and B in this study. These results are in alignment with the prevalence of flu in this state, as reported by the Georgia Department of Public Health. The baseline levels were reported as 0-3%, with an incidence of 0.6% in the second week of December 2020 (https://dph.georgia.gov/epidemiology/influenza/flu-activity-georgia).

We have previously discussed saliva samples which resulted as inconclusive and have argued that the rate of these invalid results would decrease with assays that include *RNaseP* gene as a housekeeping control [7, 12]. Herein, the number of invalids significantly decreased with the multiplex assay compared to the SARS-specific assay [1.7% (18/1019) vs. 5.6% (58/1019): p< 0.001], and Ct values for the housekeeping gene was significantly lower (p<0.001) with the multiplex assay compared to the SARS-specific assay. The *RNase P* gene is abundant in both the NPS and saliva samples, leading to a definitive result, whereas the addition of an external control (5 µl *IC* in SARS-specific assay) might be difficult to extract in complex samples contributing to inconclusive results. It is recommended that inconclusive samples be re-processed, and if the results still remain inconclusive, then patients need to be re-tested on a fresh sample. A major advantage of this multiplex assay has been that it did not cause any significant workflow changes in our laboratory. Existing platforms and even procedures remained essentially the same as those in use for the last eight months for COVID-19 testing. The only change was in the PCR master mix composition, which does not cause any change in workflow or TAT. The TAT with SARS-specific assay has been ∼14h for reporting ∼800 samples/day, and the TAT to report results with the multiplex assay was found to be similar as assessed by an independent, blinded protocol.

## Conclusion

This study demonstrates the clinical sensitivity of a novel multiplex assay and the feasibility of implementing it in laboratories without disrupting the existing laboratory workflow. However, the study is limited by the fact that none of our clinical samples were known positive for Influenza A and B viruses. To address this limitation, we obtained the SeraCare reference material for Influenza A and B viruses. We performed the LoD studies by spiking saliva and NPS samples to validate the assay for the detection of Influenza viruses. The second limitation was that Respiratory Syncytial Virus (RSV) was not validated in this study as the RT-PCR instruments available in the laboratory do not have the channel on which the dye for RSV detection was labeled. The manufacturers have appropriately provided the procedure for RT-PCR master mix preparation, where, TE (Tris-EDTA) was added instead of RSV probes in the solution. The laboratories that have instruments that can detect RSV can easily include the detection of RSV in addition to SARS-CoV-2, Influenza A, and B viruses. Nonetheless, despite these limitations, this study presents a significant and clinically validated assay that can be implemented for testing of SARS-CoV-2, Influenza A and B viruses, using NPS and saliva samples for population screening.

## Data Availability

All relevant data is provided in the manuscript.

## References

1. Taipale J, Romer P, Linnarsson S. Population-scale testing can suppress the spread of COVID- 19. medRxiv. 2020.04.27.20078329.

2. Bedford J, Enria D, Giesecke J, Heymann DL, Ihekweazu C, Kobinger G, Lane HC, Memish Z, Oh MD, Schuchat A, Ungchusak K. COVID-19: towards controlling of a pandemic. Lancet. 2020; 395(10229): 1015–8.

3. GBD 2017 Influenza Collaborators. Mortality, morbidity, and hospitalisations due to influenza lower respiratory tract infections, 2017: an analysis for the Global Burden of Disease Study 2017. Lancet Respir Med. 2019; 7(1): 69–89.

4. Xing Q, Li GJ, Xing YH, Chen T, Li WJ, Ni W, Deng K, Gao RQ, Chen CZ, Gao Y, Li Q. Precautions are needed for COVID-19 patients with co-infection of common respiratory pathogens. MedRxiv 2020; doi: https://doi.org/10.1101/2020.02.29.20027698.

5. Posteraro B, Marchetti S, Romano L, Santangelo R, Morandotti GA, Sanguinetti M, Cattani P; FPG COVID Laboratory Group. Clinical microbiology laboratory adaptation to COVID-19 emergency: experience at a large teaching hospital in Rome, Italy. Clin Microbiol Infect. 2020; 26(8): 1109–11.

6. Sahajpal NS, Mondal AK, Njau A, Ananth S, Jones K, Ahluwalia PK, Ahluwalia M, Jilani Y, Chaubey A, Hegde M, Kota V, Rojiani A, Kolhe R. Proposal of RT-PCR-Based Mass Population Screening for Severe Acute Respiratory Syndrome Coronavirus 2 (Coronavirus Disease 2019). J Mol Diagn. 2020; 22(10): 1294–99.

7. Sahajpal NS, Mondal AK, Ananth S, Njau A, Ahluwalia P, Chaubey A, Kota V, Caspary K, Ross TM, Farrell M, Shannon MP. SalivaAll: Clinical validation of a sensitive test for saliva collected in healthcare and community settings with pooling utility for SARS-CoV-2 mass surveillance. medRxiv. 2020. doi: https://doi.org/10.1101/2020.08.26.20182816.

8. Fischer WA, Ii MG, Bhagwanjee S, Sevransky J. Global burden of influenza: contributions from resource limited and low-income settings. Global heart. 2014; 9(3): 325.

9. Gordon A, Reingold A. The burden of influenza: a complex problem. Current epidemiology reports. 2018; 5(1): 1–9.

10. Bresee J, Fitzner J, Campbell H, Cohen C, Cozza V, Jara J, Krishnan A, Lee V. Progress and remaining gaps in estimating the global disease burden of influenza. Emerging infectious diseases. 2018; 24(7): 1173.

11. Lansbury L, Lim B, Baskaran V, Lim WS. Co-infections in people with COVID-19: a systematic review and meta-analysis. J Infect. 2020; 81(2): 266–275.

12. Sahajpal NS, Mondal AK, Ananth S, Njau A, Ahluwalia P, Newnam G, Lozoya-Colinas A, Hud NV, Kota V, Ross TM, Reid MD. SalivaSTAT: Direct-PCR and pooling of saliva samples collected in healthcare and community setting for SARS-CoV-2 mass surveillance. medRxiv. 2020. doi: https://doi.org/10.1101/2020.11.23.20236901.

